# A Cross-Sectional Study of COVID-19 Vaccine Hesitancy and Behaviours among People Living with HIV in British Columbia

**DOI:** 10.64898/2026.01.31.26345295

**Authors:** Anne Ejiegbu, Batoul Shariati, Jonathan Little, Mario Brondani

## Abstract

**Objective:** Although COVID-19 vaccination is important for People Living with HIV given their elevated infection and comorbidity risks, some PLHIV are hesitant to accept vaccination. Hence, we conducted a cross-sectional study in British Columbia, Canada, aimed to identify socio-economic and health-related factors predicting COVID-19 vaccine uptake and contributing to hesitancy among PLHIV.

**Methods:** A 34-item anonymous self-administered survey was disseminated to PLHIV accessing services through HIV and AIDS-related organisations’ e-newsletters between November 2022 and January 2023 in British Columbia. The survey included sociodemographic information, COVID-19 factors, HIV indicators, and the Vaccine Hesitancy Scale. Descriptive and inferential statistics were conducted to detect significant associations between the sociodemographic characteristics, health-related factors and COVID-19 vaccine uptake using IBM® SPSS® 28 and significance level at *p*<0.05.

**Results:** From the 276 respondents (mean age 29.93±7.55), 54.7% were men, 31.6% identified as sexual minorities, and 46.7% were of indigenous origin. Approximately 40% of the respondents received at least three vaccine doses, while 82.2% received at least one dose. Vaccine hesitancy was associated with lower education, age <44, and low income. Predictors of COVID-19 vaccine uptake included age [OR=1.06, 95% CI=1.01-1.12], bachelor’s degree [OR=0.22, 95% CI=0.07-0.72], family/friends infected with COVID-19 [OR 3.68 95% CI=1.56 – 8.67], HIV viral load >500 copies [OR=0.20, 95% CI=0.06-0.61], belief in vaccine importance [OR= 0.51, CI=0.28-0.95], trust in Health Canada’s information [OR 0.49 CI=0.29-0.83], and concerns about vaccine adverse effects [OR=0.35, CI=0.22-0.56]. Concerns about vaccine adverse effects reduced the likelihood of receiving three COVID-19 vaccine doses by 65%.

**Conclusions:** Considerations must be taken around specific factors that may have an impact on COVID-19 vaccination rates among PLHIV, including information about vaccine adverse effects, HIV viral load, age, and education level. This insight should guide the development of policies and interventions aimed at encouraging individuals to maintain an up-to-date vaccination status.

## Introduction

Since its first reported case, the Human Immunodeficiency Virus (HIV) has afflicted over 85.6 million people globally, leading to more than 40.4 million deaths (1). The introduction of antiretroviral therapies have substantially reduced HIV/AIDS-related morbidity and mortality. People Living with HIV (PLHIV) who adhere to therapy now attain life expectancies comparable to those without HIV, albeit mostly in developed countries (2,3).

Despite these advancements, marginalised communities, including individuals engaged in commercial sex work, men who have sex with men, people who use illicit substances, and gender minority individuals, still account for 65% of global HIV infections (4). In Canada, there are approximately 62,050 PLHIV, with about 9,800 in British Columbia (BC) (5,6).

Another global health condition came with the 2019 coronavirus disease (COVID-19) pandemic caused by the airborne SARS-CoV-2, resulting in over 704 million infections and 7.01 million deaths worldwide by December 2024 (7). Over the past four years, multiple waves of COVID-19 infections with a number of different variants have occurred globally, although there is now strong evidence that COVID-19 reached a seasonality that makes it an endemic pathogen (8). COVID-19 has also disproportionately affected marginalised communities (9). These vulnerable groups encompass PLHIV, 2SLGBTQQIA+ (Two-spirit, lesbian, gay, bisexual, trans, queer, questioning, intersex and asexual) individuals, people of colour, and Indigenous communities (10,11). Unlike the HIV epidemic’s focus on drug treatment, the COVID-19 era witnessed remarkable vaccine development. At the height of the pandemic, the Government of Canada had authorised the use of AstraZeneca (Vaxzevria), Janssen (Johnson & Johnson), Medicago (Covifenz), Moderna (Spikevax), Novavax (Nuvaxovid), and Pfizer-BioNTech (Comirnaty) vaccines (12). Vaccination programmes primarily aim to prevent and control infectious diseases, ultimately striving for herd immunity and reducing person-to-person transmission (13). Initial studies suggested that achieving herd immunity against COVID-19 would necessitate vaccinating 60-70% of the population using vaccines with 70-90% efficacy (14). However, the still ongoing mutations in the SARS-CoV-2 virus have challenged these estimates (15).

During the initial phases of the COVID-19 pandemic, concerns arose regarding the heightened vulnerability of PLHIV to severity of the disease (16). However, estimates reveal that PLHIV constituted approximately 1.0% of hospitalised cases at that time, with the prevalence of SARS-CoV-2 infection ranging from 0.88% to 1.8% (17,18), analogous to that of the general population (19). While PLHIV were not overrepresented in COVID-19 cases, they experienced more severe outcomes attributable to HIV-related social disparities, medical risks, and comorbidities (20,21). Available data indicate vaccine efficacy and safety among individuals with well-managed HIV, undetectable viral loads, and CD4 cell counts exceeding 200 cells/mm^3^ (30-22).

In the context of COVID-19 immunisations, “full vaccination” refers to completing at least the recommended primary series of vaccines (23). Conversely, being ‘up to date’ with COVID-19 vaccination implied that an individual had received all the recommended vaccine series doses, plus the two boosters by earlier 2023 (24,25). Multiple additional doses (boosters) continue to be required given the COVID-19 mutations and enhanced transmissibility (26). Boosters are suggested, but not without increasing uncertainties and vaccine fatigue (27) even when offered along with other seasonal vaccination regimes such as the annual flu vaccines (28). Nonetheless, the success of vaccination programs hinges on adherence to these public health recommendations (29). Vaccine uptake rate signifies the proportion of the population who have received a vaccine within a specific period, and is influenced by acceptance, hesitancy, and refusal. Vaccine hesitancy is defined as delayed approval, or rejection altogether of vaccines (29), and is influenced by complacency, convenience, and confidence (30). Hesitancy is compounded by deep-seated mistrust in the medical system, adherence to public health directives (31), and the anti-vax movement and conspiracy theories (32). For individuals living with HIV, additional apprehensions include fear of contracting COVID-19 from the vaccines and potential effects (33). Despite numerous research in Canada investigating PLHIV’s willingness to receive COVID-19 vaccinations during the initial years of the pandemic (34,35), there was limited information at that time on the actual uptake and adherence to the recommended primary vaccine series. Furthermore, there was limited research exploring the intricate interplay between vaccine uptake and hesitancy and health-related factors, including HIV indicators such as CD4+ count, HIV viral load, comorbidities, length of HIV diagnosis, perceived vulnerability, socioeconomic considerations, and prevailing social norms among PLHIV. Given this paucity of literature, this study addressed the following research question ‘*What are the socio-economic and health-related predicting factors of COVID-19 uptake and associated factors of hesitancy among PLHIV in British Columbia, Canada?*’ aided by three objectives: to determine the prevalence of COVID-19 vaccine uptake among PLHIV in British Columbia, Canada; the level of vaccine hesitancy among PLHIV in British Columbia using the adult vaccine hesitancy scale; and the socio-economic and health (COVID-19 and HIV-related) factors associated with COVID-19 vaccine uptake (including predictors) and vaccine hesitancy within 2022 and 2023.

## Methods

### Study Design and Population

This study adopted a cross-sectional design via a 34-item survey pilot tested incorporating the Adult Vaccine Hesitancy Scale - VHS (36). The self-administered survey was conducted anonymously via Qualtrics© software (2019, Provo, Utah). Participation was voluntary and anonymous; informed consent was obtained through the introductory page.

The survey consisted of questions about demographics (age, gender, ethnicity, etc); COVID-19 vaccination status (number of doses, motivation and deterrents to vaccination); perceived COVID-19 vulnerability (previous COVID-19 diagnosis, family & friend diagnosis and/or death from COVID-19); HIV health indicators (CD4 count, HIV viral load), and VHS as validated by Akel et al (36) with 10 Likert scale questions with agreement levels (strongly agree to strongly disagree) with statements like ‘Vaccines are important for my health’, and ‘Vaccines are effective’. Scores in each statement range from 1 to 5, totalling 10 (minimum) and 50 (maximum).

Inclusion criteria encompassed adults 19 years of age or older, of any gender or sexual orientation, who self-reported a diagnosis of HIV/AIDS during data collection, were willing to participate and able to understand English to fill out the survey. Exclusion Criteria encompassed anyone younger than 19 years living or not with HIV, and residing outside British Columbia.

#### Sample Size Determination

Sample size calculation using the Cochran formula set the minimal sample to 384, assuming a normal standard deviation set at 1.96, corresponding to a 95% confidence level, a degree of variability set at a maximum 50%= 0.50 and a level of precision/ margin of error set at ±0.05 (5%). This study employed a convenience sampling method in collaboration with HIV organisations in British Columbia that distributed the survey link to their members between the 17th of October 2022 and the 7th of January 2023.

### Data Analyses

Data was analysed using IBM® SPSS® 28 for Mac (SPSS Inc., Chicago, IL, USA, 2021). Descriptive statistics was employed to summarise socio-demographic characteristics. Inferential statistics used two outcome variables, including total VHS, and COVID-19 Vaccine Uptake (comprising at least three doses + booster doses). The Shapiro-Wilk’s W-test yielded a non-significant result (p=0.080), confirming the fulfilment of the normality. Pearson’s Product-Moment Correlation and one-way Analysis of Variance (ANOVA) were then performed. For the COVID-19 Vaccine Uptake, answers were dichotomised into YES (received primary doses + boosters) and NO (did not receive primary doses + boosters). Bivariate analysis was done using Pearson’s Chi-square test for independence after fulfilling the lowest expected cell frequency assumption (>25%). Variables were significant at p<0.05. Multivariate analysis was carried out using binary logistic regression. The independent variables included in the model were those with p<0.10 in addition to those with a significant p<0.05 from the bivariate analysis. The model was significant using the Omnibus Tests of Model Coefficients (χ2 = 98.36; df = 27; p<0.001; classification fit =76.7%) and non-significant

## Results

A total of 477 responses were from British Columbia; 276 were PLHIV with a mean age at the time of data collection of 29.93 [SD = 7.55; range, 19–67 years], from a diverse ethnic and demographic background with men comprising 54.7% of participants, with sexual minorities making up 31.6%, and with Indigenous individuals accounting for 46.7% of the total population, as shown in **Table 1**.

As of January 2023, completing the primary series meant receiving the recommended three doses, while being up-to-date entailed having two recommended boosters, totalling 5 doses. Partial vaccination was defined as receiving less than 5 doses.

**Table 1:**
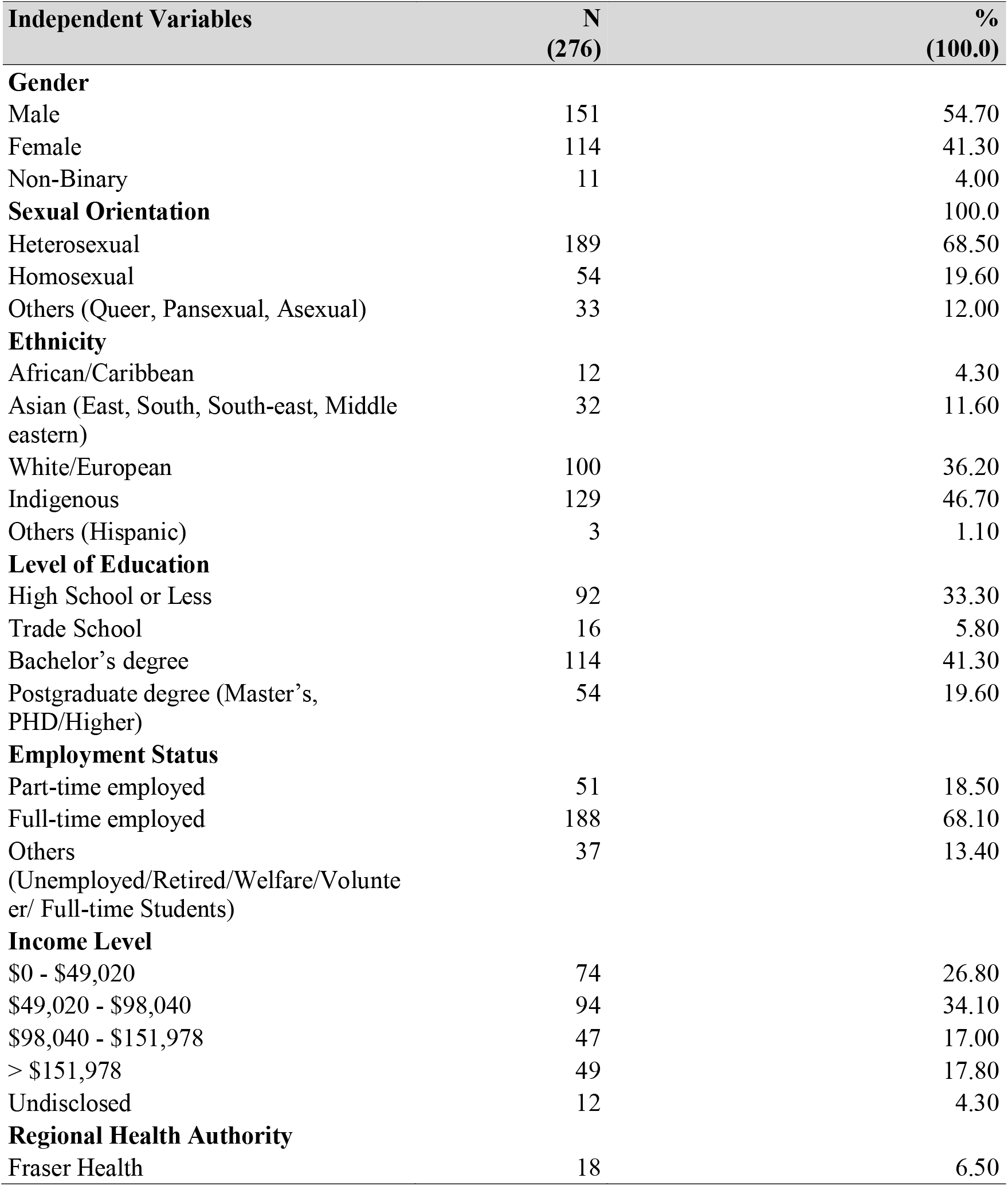
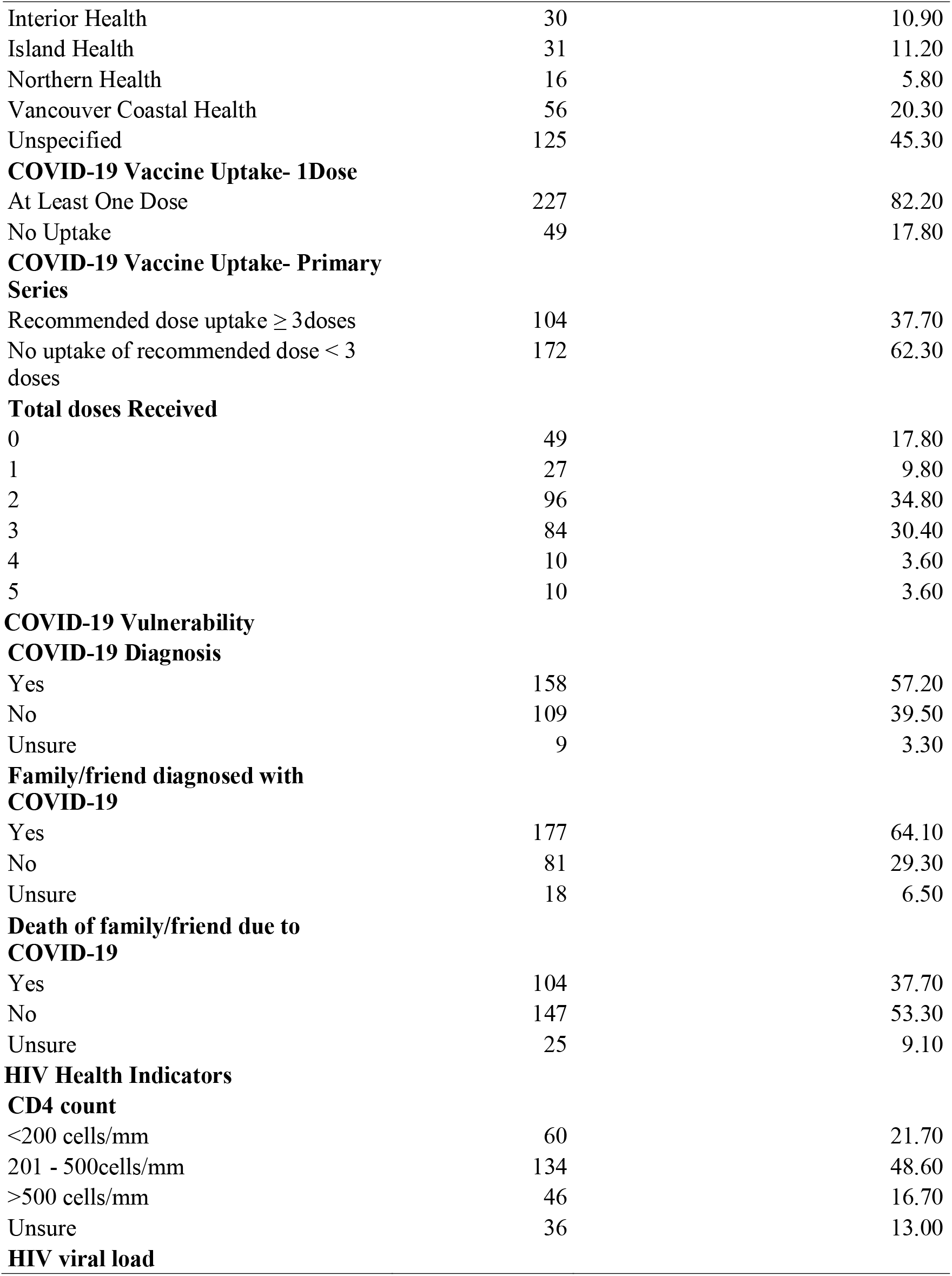
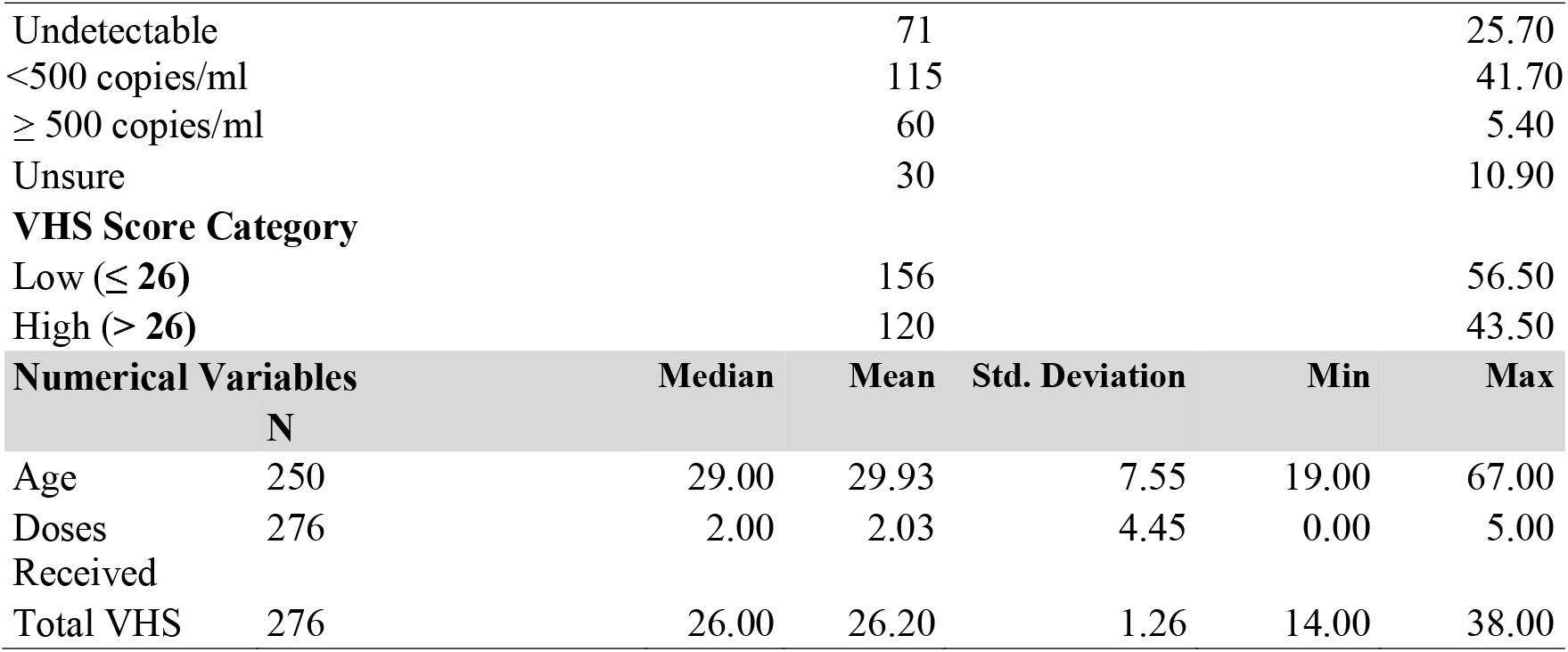
Sociodemographic Profile of the 276 PLHIV in British Columbia who Participated in the Study.

| **Independent Variables** | **N**<br>**(276)** | **%**<br>**(100.0)** |
| --- | --- | --- |
| **Gender** |  |  |
| Male | 151 | 54.70 |
| Female | 114 | 41.30 |
| Non-Binary | 11 | 4.00 |
| **Sexual Orientation** |  | 100.0 |
| Heterosexual | 189 | 68.50 |
| Homosexual | 54 | 19.60 |
| Others (Queer, Pansexual, Asexual) | 33 | 12.00 |
| **Ethnicity** |  |  |
| African/Caribbean | 12 | 4.30 |
| Asian (East, South, South-east, Middle eastern) | 32 | 11.60 |
| White/European | 100 | 36.20 |
| Indigenous | 129 | 46.70 |
| Others (Hispanic) | 3 | 1.10 |
| **Level of Education** |  |  |
| High School or Less | 92 | 33.30 |
| Trade School | 16 | 5.80 |
| Bachelor's degree | 114 | 41.30 |
| Postgraduate degree (Master's, PHD/Higher) | 54 | 19.60 |
| **Employment Status** |  |  |
| Part-time employed | 51 | 18.50 |
| Full-time employed | 188 | 68.10 |
| Others<br>(Unemployed/Retired/Welfare/Volunteer/ Full-time Students) | 37 | 13.40 |
| **Income Level** |  |  |
| \$0 - \$49,020 | 74 | 26.80 |
| \$49,020 - \$98,040 | 94 | 34.10 |
| \$98,040 - \$151,978 | 47 | 17.00 |
| > \$151,978 | 49 | 17.80 |
| Undisclosed | 12 | 4.30 |
| **Regional Health Authority** |  |  |
| Fraser Health | 18 | 6.50 |
| Interior Health | 30 | 10.90 |
| Island Health | 31 | 11.20 |
| Northern Health | 16 | 5.80 |
| Vancouver Coastal Health | 56 | 20.30 |
| Unspecified | 125 | 45.30 |
| **COVID-19 Vaccine Uptake- 1Dose** |  |  |
| At Least One Dose | 227 | 82.20 |
| No Uptake | 49 | 17.80 |
| **COVID-19 Vaccine Uptake- Primary Series** |  |  |
| Recommended dose uptake $\geq 3$ doses | 104 | 37.70 |
| No uptake of recommended dose $< 3$ doses | 172 | 62.30 |
| **Total doses Received** |  |  |
| 0 | 49 | 17.80 |
| 1 | 27 | 9.80 |
| 2 | 96 | 34.80 |
| 3 | 84 | 30.40 |
| 4 | 10 | 3.60 |
| 5 | 10 | 3.60 |
| **COVID-19 Vulnerability** |  |  |
| **COVID-19 Diagnosis** |  |  |
| Yes | 158 | 57.20 |
| No | 109 | 39.50 |
| Unsure | 9 | 3.30 |
| **Family/friend diagnosed with COVID-19** |  |  |
| Yes | 177 | 64.10 |
| No | 81 | 29.30 |
| Unsure | 18 | 6.50 |
| **Death of family/friend due to COVID-19** |  |  |
| Yes | 104 | 37.70 |
| No | 147 | 53.30 |
| Unsure | 25 | 9.10 |
| **HIV Health Indicators** |  |  |
| **CD4 count** |  |  |
| <200 cells/mm | 60 | 21.70 |
| 201 - 500cells/mm | 134 | 48.60 |
| >500 cells/mm | 46 | 16.70 |
| Unsure | 36 | 13.00 |
| **HIV viral load** |  |  |

|  |  |  |  |  |  |  |
| --- | --- | --- | --- | --- | --- | --- |
| Undetectable |  | 71 |  |  |  | 25.70 |
| <500 copies/ml |  | 115 |  |  |  | 41.70 |
| ≥ 500 copies/ml |  | 60 |  |  |  | 5.40 |
| Unsure |  | 30 |  |  |  | 10.90 |
| **VHS Score Category** |  |  |  |  |  |  |
| Low (≤ 26) |  | 156 |  |  |  | 56.50 |
| High (> 26) |  | 120 |  |  |  | 43.50 |
| **Numerical Variables** |  | **Median** | **Mean** | **Std. Deviation** | **Min** | **Max** |
|  | **N** |  |  |  |  |  |
| Age | 250 | 29.00 | 29.93 | 7.55 | 19.00 | 67.00 |
| Doses Received | 276 | 2.00 | 2.03 | 4.45 | 0.00 | 5.00 |
| Total VHS | 276 | 26.00 | 26.20 | 1.26 | 14.00 | 38.00 |

### COVID-19 Vaccine Hesitancy

The mean vaccine hesitancy score was 26.20± 1.260 and was negatively correlated with age [r= –.18, n=250, p<.0005]. There was also a negative correlation between the number of doses received and the vaccine hesitancy score value [r= –.245, n=276, p<.001]. One-way ANOVA revealed a statistically significant difference in the mean VHS scores between those younger than 25 and those between 26 and 44 years old [F (3, 272) =7.18, p<0.001]. Post-hoc comparisons using the Turkey HSD test indicated the mean score for participants aged ≤25 (M=27.04, SD=4.34) had significantly different scores from those aged 26-44 (M=25.32, SD=4.28) (p = 0.019, 95% C.I. = [0.21, 3.22]). Age ≥45 (M=25.20, SD=5.16) did not significantly differ from either younger age group.

Similarly, there was a significant effect of the level of education on VHS scores [F (3, 272) =6.42, p<0.001]. Participants with a bachelor’s degree (M=24.86, SD=0.39) had significantly lower mean scores than those with a high school education or less (M=26.92, SD=0.47) (p = 0.004, 95% C.I. = [-3.63, -0.50]).

Income level was also statistically significant [F (4, 271) =5.00, p<0.001]. Participants in the lowest federal tax bracket earning ≤CD$49,020 (M=24.68, SD=4.43) had mean VHS scores that were significantly lower than those who earned CD$98,040-CD$151,978 (M=27.00, SD=3.90)-(p = 0.03, 95% C.I. = [-4.54, -0.11]).

Lastly, although HIV viral load revealed a significant output [F (3, 272) =2.82, p=0.04], the post-hoc comparisons revealed no statistically significant difference between either group of viral loads, i.e., undetectable, <500 Copies or ≥500 copies.

### COVID-19 Vaccine Uptake

Approximately 82.2% of the participants had received at least one dose of the COVID-19 vaccine, while 17.8% (n=49) remained unvaccinated at the time of this study. Around 37.6% (n = 104) were vaccinated with at least the recommended primary series of three doses, while 3.6% (n = 10) were up to date as of January 2023, having received all five recommended doses. Notably, more than half of the participants had received a COVID-19 diagnosis (n = 158, 59.2%) or were aware of a positive COVID-19 diagnosis among family or friends (n = 177, 64.1%).

Multi-variable modelling in the form of binary logistic regression analyses was utilised to explain COVID-19 vaccine uptake (**Table 2**); the model was significant using the Omnibus Tests of Model Coefficients (χ2 = 98.36; df = 27; p<0.001; classification fit =76.7%) and non-significant (χ2 = 12.77; df=8; p=.120) using Hosmer and Lemeshow test, both indicating a good model fit.

The model exhibited a sensitivity of 62.5% for correctly identifying individuals vaccinated with the recommended primary series and a specificity of 85.6%. Furthermore, the model’s positive predictive value was 73.2%, and its negative predictive value stood at 78.4%. The likelihood of being vaccinated with at least 3 doses increased by 1.06 [95% CI-1.01 –1.12, p= 0.019] as the age of the participants increased by one year. Conversely, participants with bachelor’s degrees had 78% lower odds [95% CI-0.07 - 0.72, p= 0.012] of completing the recommended 3 doses or more compared to those with high school education. Moreover, participants with friends or family having a COVID-19 history had a 3.68 increased likelihood of completing the recommended 3 doses or more [95% CI 1.56 – 8.67, p = 0.003]. Additionally, participants with a viral load of less than 500 copies/ml had 80% lower odds of receiving at least 3 doses compared to those with undetectable viral loads [95% CI 0.01 - 0.81, p = 0.019]. Total vaccine hesitancy score was seen to be a predictor of vaccine uptake; paradoxically, an increase in one unit of the total vaccine hesitancy score was seen to increase the likelihood of receiving the recommended doses by 1.35 [95% CI= 1.06 - 1.73, p= 0.015]. However, a breakdown of the individual scores on the scale revealed that an increase in scores reflecting disagreement about vaccine importance, doubts about Health Canada’s information reliability, and concerns about serious adverse effects led to reduced odds of receiving the recommended ≥ 3 COVID-19 vaccine doses by 48% [95% CI 0.28 - 0.95, p = 0.015], 51% [95% CI 0.29 - 0.84, p = 0.008], and 65% [95% CI 0.22 - 0.56, p < 0.001], respectively.

**Table 2:** Logistic Regression Models Analyzing COVID-19 Vaccine Uptake in PLHIV in British Columbia.

|  | Received At Least Three Doses Of COVID-19 Vaccine (N=276) |  |  |  |  |
| --- | --- | --- | --- | --- | --- |
|  | **B** | **Wald** | **OR** | **95% CI** | **p** |
| <u>Age</u> | 0.06 | 5.51 | 1.06 | **1.01 – 1.12** | **0.019** |
| <u>Level of Education</u> |  | 6.75 |  |  | 0.080 |
| High School or Less | -0.77 | 0.90 | 0.46 | 0.09 - 2.27 | 0.343 |
| Trade School | -0.35 | 0.73 | 0.70 | 0.32 – 1.58 | 0.392 |
| Bachelor's Degree | -1.54 | 6.29 | 0.22 | **0.07 - 0.72** | **0.012** |
| <u>Employment status</u> |  | 2.27 |  |  | 0.322 |
| Part-time employed | 0.01 | 0.00 | 0.99 | 0.26 – 3.78 | 0.986 |
| Full-time employed | -0.64 | 1.03 | 0.53 | 0.16 – 1.81 | 0.311 |
| <u>Income Level</u> |  | 7.09 |  |  | 0.131 |
| \$0 - \$49,020 | -0.25 | 0.27 | 0.78 | 0.31 - 1.98 | 0.601 |
| \$49,020 - \$98,040 | 0.57 | 0.94 | 1.77 | 0.56 - 5.61 | 0.333 |
| \$98,040 - \$151,978 | -0.88 | 1.78 | 0.42 | 0.12 - 1.51 | 0.183 |
| > \$151,978 | 0.97 | 1.01 | 2.64 | 0.40 - 17.50 | 0.314 |
| <u>Family/friend diagnosed of COVID 19</u> |  | 11.60 |  |  | **0.003** |
| Family/friend diagnosed of COVID 19 (Yes) | 1.30 | 8.87 | 3.68 | **1.56 – 8.67** | **0.003** |
| Family/friend diagnosed of COVID 19 (No) | -1.12 | 1.15 | 0.33 | 0.04 – 2.53 | 0.283 |
| <u>HIV Viral Load</u> |  | 14.90 |  |  | **0.002** |
| Undetectable | 0.51 | 0.95 | 1.67 | 0.60 - 4.69 | 0.330 |
| <500 copies/ml | -1.63 | 8.05 | 0.20 | **0.06 - 0.61** | **0.005** |
| ≥ 500 copies/ml | -0.76 | 0.96 | 0.47 | 0.10 - 2.22 | 0.327 |
| <u>CD4+ Count</u> |  | 3.26 |  |  | 0.353 |
| <200 cells/mm | 0.46 | 0.76 | 1.58 | 0.56 - 4.43 | 0.330 |
| 201 - 500cells/mm | 1.03 | 3.04 | 2.79 | 0.88 – 8.86 | 0.081 |
| >500 cells/mm | 0.80 | 1.01 | 2.22 | 0.47 - 10.54 | 0.315 |
| Total VHS | 0.30 | 5.95 | 1.35 | **1.06 - 1.73** | **0.015** |
| Vaccines are important for my health | -0.66 | 4.48 | 0.52 | **0.28 - 0.95** | **0.034** |
| Vaccines are effective | -0.13 | 0.20 | 0.88 | 0.51 - 1.53 | 0.655 |
| Being vaccinated is important for the health of others in my community. | -0.42 | 1.88 | 0.65 | 0.36 - 1.20 | 0.170 |
| All routine vaccinations recommended by Health Canada are beneficial | -0.10 | 0.10 | 0.91 | 0.50 - 1.65 | 0.747 |
| New vaccines carry more risks than older vaccines. | 0.06 | 0.06 | 1.06 | 0.68 - 1.65 | 0.809 |
| The information I receive about vaccines from Health Canada is reliable and trustworthy | -0.71 | 6.98 | 0.49 | **0.29 - 0.83** | **0.008** |
| Generally, I do what my doctor or healthcare provider recommends about vaccines for me | -0.58 | 3.37 | 0.56 | 0.30 - 1.04 | 0.066 |
| I am concerned about serious adverse effects of vaccines | -1.05 | 19.58 | 0.35 | **0.22 – 0.56** | **<0.001** |

## Discussion

This study determined the prevalence of COVID-19 vaccine uptake and level of vaccine hesitancy among PLHIV in British Columbia, Canada, during winter of 2023. It also focused on the socio-economic and health (COVID-19 and HIV-related) factors associated with COVID-19 vaccine uptake and vaccine hesitancy at a time when COVID vaccines (and boosters) still had a fair uptake compared to the current 2024/25 winter season (37). Our findings expand our understanding of COVID-19 vaccine uptake and hesitancy among PLHIV.

While the initial vaccine uptake of at least one dose mirrored the general population at 82.2% vs 83.3%, only 37.4% of PLHIV in our study completed the primary vaccine series—significantly lower than the national average (80.6% as of January 2023) (38). This discrepancy is unsurprising given that earlier studies demonstrated significantly lower rates than the general population, stemming from reduced confidence in vaccination and specific concerns unaddressed by provincial interventions in this demographic (39). A recurring theme among PLHIV who refused or only partially received vaccines was a lack of sufficient information and mistrust in Health Canada’s guidance. This underscores the importance of tailored communications catering to the unique social and medical circumstances of this population (40,41). Moreover, limited PLHIV were included in the vaccine trials, which might have further influenced any scepticism against the results of those trials for PLHIV. It would be of interest to repeat the study, or conduct a follow up with previous participants, during this current winter of 2025 to explore the extent to which the combined offering of both COVID-19 booster and seasonal flu vaccines influences uptake. Of note, older individuals completed the primary series, while younger ones displayed higher vaccine hesitancy scores, as seen elsewhere (40,42). This emphasises the need for targeted health promotion strategies to engage younger individuals, many of whom perceive COVID-19 to be of lower risk and severity.

Contrary to the prevailing trend of those with higher education having an increased vaccine acceptance (43), our study showed a negative association between having a bachelor’s degree and vaccine uptake among PLHIV. This finding may suggest that hesitancy cannot solely be attributed to educational attainment or may be due to the fact that our sample size was below the minimal estimated to be significant. While educational level can serve as a valuable indicator of access to reliable health information sources, it is crucial to recognise the profound impact of misinformation and disinformation on pandemic responses globally (44,45) specially amidst the new variants of SARS-CoV-2 (46).

Regarding the income level, participants in the lowest earning bracket had a higher mean vaccine hesitancy score, corroborating other studies, albeit not with PLHIV (40,42). Still, income level, sexual orientation, gender and ethnicity were not predicting factors of uptake of three doses. This may be due to the small number of participants.Considering HIV-related factors, only HIV viral load was seen to be related to uptake, where having a viral load of 500 or less decreased the odds of receiving the recommended three doses compared to those with undetectable viral loads, as also found elsewhere (45). Perception of vulnerability to the COVID-19 disease is also worth considering, as our study highlighted participants who had family or friends with a history of positive COVID-19 diagnosis had higher likelihood of completing the recommended three doses plus boosters. This may increase awareness of the disease and the natural inclination to prevent being infected (38).

### Strengths and Limitations

Our study revealed that a high vaccine hesitancy score did not hinder receiving the recommended doses. Regardless, concerns about potential adverse effects and lack of trust in Health Canada’s information, combined with doubts about vaccine benefits on health, exert the most substantial influence on vaccine uptake. These concerns outweigh the overall hesitancy score, indicating that individuals with lower scores can still exhibit pronounced hesitancy if they firmly hold these beliefs. Health promotion campaigns targeting highly hesitant populations should be well-rounded in their considerations.

Despite its findings, our study is not without limitations. The survey developed was not validated following psychometric assessment (47), although it was piloted and modified accordingly. The number of participants below the minimal sample size prevents generalisation. As with any self-reported data, findings are subject to social desirability bias. Additionally, this cross-sectional study precludes the establishment of causality. Furthermore, electronic data collection excluded those who lack access to digital channels. Lastly, it is vital to acknowledge the temporal limitations of our data as the evolving nature of the COVID-19 pandemic render our findings specific to the winter of 2023. Nevertheless, these findings can serve as a valuable reference point for future investigations into emerging trends related to COVID-19 and vaccine acceptance.

Albeit the acknowledged limitations, our research provides a thorough examination of COVID-19 vaccine uptake by PLHIV. The insights gleaned from our study underscore the need for a more nationally representative investigation across BC and Canada. Such an expanded study should also monitor up-to-date dose uptake over specific timeframes to discern evolving trends and validate predictive factors.

## Conclusion

Our findings reinforce the idea of continuously considering specific factors that may have an impact on vaccination rates, including information about adverse effects and the importance of vaccines among PLHIV. The findings of our study have also revealed that vaccination hesitancy and uptake of one vaccine dose are subject to change and do not necessarily predict the uptake of the necessary additional doses in the case of this highly mutagenic SARS-COv2 virus. Hence, aside from socioeconomic factors, perceptions regarding HIV-related factors should be addressed within this demographic. This insight can further guide the development of policies and interventions aimed at promoting additional vaccine doses and encouraging individuals to maintain an up-to-date vaccination status.

## Data Availability

All data produced in the present study are available upon reasonable request to the authors.

## Ethics approval

Ethical approval for this study was granted by the Behavioral Research Ethics Board at the University of British Columbia (UBC) # H22-02053 in accordance with the Declaration of Helsinki.

## Consent to participate

Consent to participate was obtained in written by the participants via the informed consent found at https://open.library.ubc.ca/soa/cIRcle/collections/ubctheses/24/items/1.0437303, page 88.

## Consent for publication

Consent for publication was obtained in written by the participants via the informed consent found at https://open.library.ubc.ca/soa/cIRcle/collections/ubctheses/24/items/1.0437303, page 88.

## Availability of data and materials

Tabulated data from the surveys can be made available upon request to the corresponding author. The survey used for this study is presented as supplementary information file. The UBC Office of Research Ethics requires safeguards on our part, so only selected transcripts would be shared once clearance is given and upon request (Tel: 604-822-8598 or), and provided the transcript meet the criteria for access to confidential data.

## Competing interests

The authors declare they have no competing or conflict of interest.

## Funding

Not applicable

## Authors’ contributions

A Ejiegbu and M Brondani made equal contributions to the manuscript, as M Brondani was the supervisory professor through the process, including conceptualisation, methodology and writing. Conceptualisation, proposal, and review were carried out by A Ejiegbu, B Shariati, J Little and M Brondani, and then data analysis was carried out by A Ejiegbu and supervised by B Shariati.

## Acknowledgements

The authors gratefully acknowledge the valuable collaboration of the Pacific AIDS Network AIDS Vancouver and Community Based Research Centre. Additionally, this study’s findings were presented at the 2023 International Association of Dental Research Conference, 101^st^ General Session, in Bogota, Colombia. This article is based on a thesis submitted for partial fulfillment of the requirements of AE from the University of British Columbia for the degree of Master of Science in Craniofacial Sciences, successfully defended in October 2023.

## Notes

### Competing Interest Statement

The authors have declared no competing interest.

### Funding Statement

This study did not receive any funding.

### Author Declarations

Ethical approval for this study was granted by the Behavioral Research Ethics Board at the University of British Columbia H22-02053.

